# The impact of climate change and natural climate variability on the global distribution of *Aedes aegypti*

**DOI:** 10.1101/2023.08.31.23294902

**Authors:** AR Kaye, U Obolski, L Sun, JW Hurrell, MJ Tildesley, RN Thompson

## Abstract

*Aedes aegypti* spread pathogens affecting humans, including the dengue, Zika and yellow fever viruses. Anthropogenic climate change is altering the spatial distribution of *Ae. aegypti* and therefore the locations at risk of vector-borne disease. In addition to climate change, natural climate variability, resulting from internal atmospheric processes and interactions between climate system components (e.g. atmosphere-land, atmosphere-ocean) determines climate outcomes. However, the combined effects of climate change and natural climate variability on future *Ae. aegypti* spread have not been assessed fully. We developed an ecological model in which *Ae. aegypti* population dynamics depend on climate variables (temperature and rainfall). We used 100 projections from the Community Earth System Model, a comprehensive climate model that simulates natural climate variability as well as anthropogenic climate change, in combination with our ecological model to generate a range of equally plausible scenarios describing the global distribution of suitable conditions for *Ae. aegypti* up to 2100. Like other studies, we project the poleward expansion of *Ae. aegypti* under climate change. However, the extent of spread varies considerably between projections, each under the same Shared Socioeconomic Pathway scenario (SSP3-7.0). For example, by 2100, climatic conditions in London may be suitable for *Ae. aegypti* for between one and five months in the year, depending on natural climate variability. Our results demonstrate that natural climate variability yields different possible future *Ae. aegypti* spread scenarios. This affects vector-borne disease risks, including the potential for some regions to experience outbreaks earlier than expected under climate change alone.

## Introduction

Climate-sensitive infectious diseases pose a substantial threat to public health.^1^ Anticipating the locations in which outbreaks are most likely to occur in future allows limited surveillance resources to be deployed effectively.

Vector-borne diseases, and their vectors such as *Ae. aegypti*, are particularly sensitive to climate variations.^2^ Climate variables such as temperature and rainfall affect vector ecology, including the vector lifespan,^3^ the probability of egg survival^4, 5^ and the development time from eggs to adults.^6^ Laboratory-based research has been complemented by observational capture-release studies^7, 8^ and modelling analyses,^9–12^ demonstrating that the impacts of climate on vector dynamics are substantial and complex. Projecting the effects of future climate on vector-borne disease outbreaks therefore requires an improved understanding of the relationship between climate and the ecology of vectors such as *Ae. aegypti*.

Mathematical models have been developed and applied to study the impact of anthropogenic climate change on the spatial distribution of mosquitoes. These studies have considered different future greenhouse gas concentration trajectories and variability in projections driven by different climate models.^13, 14^ However, these represent only two of the potential sources of uncertainty in projections of future climate.^15^ In addition to anthropogenic climate change, Earth’s future climate trajectory will be strongly influenced by natural climate variability.

Natural (or internal) climate variability^15–17^ refers to those fluctuations in climate that occur even if there are no changes in the radiative (“external”) forcing of the planet. Quantitative estimates of the role of natural climate variability relative to the forced anthropogenic climate change signal can be obtained from an ensemble of projections generated by the same climate model. Each simulation, subjected to the same external forcing but initiated from slightly different initial states, will diverge due to the chaotic dynamics of the climate system. Despite the importance of natural climate variability in projections of future climate, a thorough investigation into the combined effects of anthropogenic climate change and natural climate variability on vector ecological dynamics has not previously been undertaken.

Here, we address this and develop a mathematical modelling framework for projecting the impacts of climate change and natural climate variability on the global distribution of suitable climate conditions for *Ae. aegypti*. To do this, it is necessary to use a climate projection model that can generate a large ensemble of forward climate projections. We consider 100 projections of global temperature and rainfall from the Community Earth System Model^18–20^ (CESM), a primary climate simulation model in the USA. Each projection is run under the same Shared Socioeconomic Pathway scenario (SSP3-7.0) but is initiated from a slightly different initial state. Thus, the climate conditions at any future time can be thought of as a random sample from the CESM projections at that time.

We use the CESM projections as inputs to an ecological model in which *Ae. aegypti* population dynamics depend on temperature and rainfall. We investigate the regions that might be expected to experience an expansion in suitable conditions for *Ae. aegypti* over the remainder of the 21^st^ century. We explore the impact of natural climate variability by assessing the variation in the number of months that are suitable for *Ae. aegypti* each year between different climate projections. This research represents the first detailed study of the combined effects of climate change and natural climate variability on the temporal and global spatial distribution of environmental suitability for *Ae. aegypti*, with implications for future vector-borne disease outbreak risks.

## Methods

### Climate projections

Climate data were obtained from the CESM version 2 Large Ensemble Community Project (LENS2).^20^ The dataset consists of 100 equally plausible climate simulations run from 1850 to 2100, with a range of different oceanic and atmospheric initial states between simulations. The projected impact of the externally forced anthropogenic climate change signal can be obtained by averaging over all 100 of the climate simulations. Real-world climate dynamics are far less smooth than this ensemble mean (Fig S1), illustrating the need to consider natural climate variability in addition to anthropogenic climate change when inferring the effects of future climate on *Ae. aegypti* populations.

CESM projections assume that socioeconomic changes up to the year 2100 occur under the Shared Socioeconomic Pathway 3 (SSP3-7.0) scenario. The CESM LENS2 dataset provides the values of projected climate variables across a global grid consisting of 192 latitude and 288 longitude values. In our main analyses, we consider the average temperature and rainfall each month in each location and each CESM simulation individually in the period from 2020 to 2100. Since the parameters of the ecological model (see below) depend on daily rainfall rather than monthly rainfall, we then convert the average monthly rainfall to its corresponding daily value.

### Ecological model

Temporal *Ae. aegypti* dynamics in each location are modelled using a compartmental, ordinary differential equation model. Members of the *Ae. aegypti* population are divided according to their life cycle stage: eggs, aquatic (larvae or pupae) and adults (Fig 1A). Each model parameter, including the rates at which vectors move between these stages, is assumed to depend on either temperature or rainfall.

**Figure 1.**
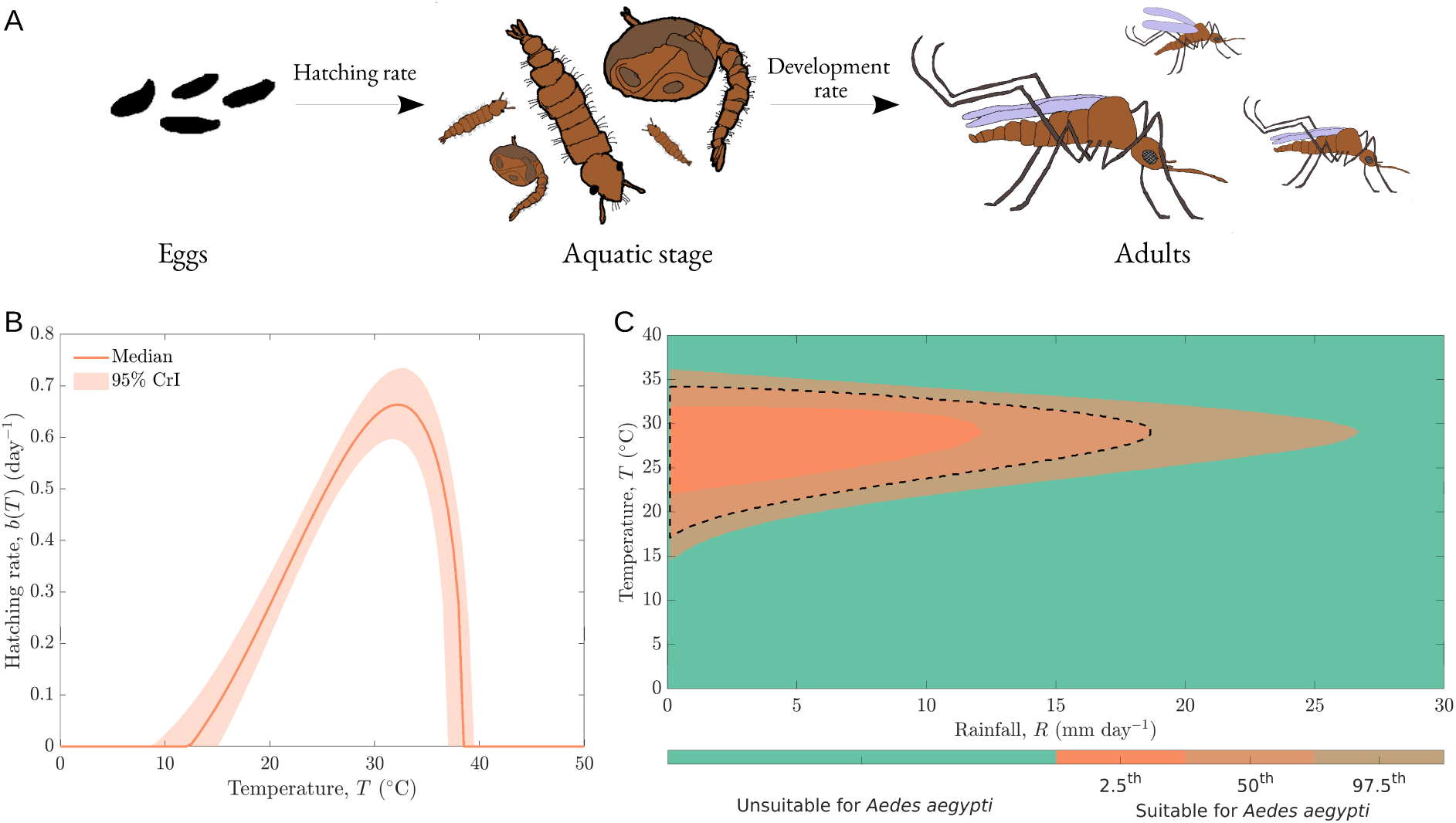
The climate-sensitive model of *Ae. aegypti* population dynamics. A. Schematic illustrating the life cycle of *Ae. aegypti*; each stage is represented by a different compartment in the ecological model. B. Ecological parameters are assumed to depend on either temperature or rainfall. Here, the dependence of the *Ae. aegypti* egg hatching rate on temperature is shown, estimated using data from a previous study^21^ (median estimate – red; 95% credible interval (CrI) – shaded region). C. The ecological niche, describing temperature-rainfall values at which an *Ae. aegypti* population can be sustained (orange), as derived from the ecological model. Uncertainty in the ecological niche is represented by different shades of orange (representing the 2.5^th^, 50^th^ and 97.5^th^ percentile estimates) and arises due to uncertainty in the parameter estimates of the ecological model. The black dotted line indicates the outline of the median ecological niche.

To infer the relationship between temperature and each temperature-dependent model parameter, we use the modelling framework of Mordecai *et al.*^21^ In brief, a general functional form of the relationship between temperature and each model parameter is assumed. We fit the precise temperature-dependent response to data from that study using Markov chain Monte Carlo (MCMC). As an example, the dependence of the egg hatching rate (the rate at which eggs hatch and enter the aquatic stage) on temperature is shown in Fig 1B. Since we adopt a Bayesian approach, we obtain a range of plausible temperature-dependent responses for each parameter (corresponding to different steps of the MCMC chain). In Fig 1B, the median egg hatching rate at each temperature value is shown in red, with the shaded area representing the 95% credible interval (CrI). Further details about the ecological model are given in the Supplementary Material. Results from the model fitting procedure are shown in Figs S2-S6 and Tables S1 and S2, with responses analogous to Fig 1B but for all temperature-dependent model parameters shown in Fig S6A-D. Two model parameters are assumed to depend on rainfall. The aquatic stage carrying capacity is assumed to increase with higher rainfall. In contrast, survival of aquatic stage individuals decreases with higher rainfall, as larvae may be washed away. Relationships between rainfall and these model parameters are derived from first principles using a mechanistic approach (Supplementary Material and Fig S6E,F).

## Results

### Ecological niche

To begin to understand the impact of temperature and rainfall on *Ae. aegypti* survivability, we first considered the combinations of temperature-rainfall values at which the ecological model predicts *Ae. aegypti* can survive (the ecological niche). Specifically, we calculated the equilibrium population size of the ecological model at each possible temperature-rainfall combination, and indicated the values at which the equilibrium population size is greater than zero so that an *Ae. aegypti* population can be supported (Fig 1C). Uncertainty in the precise dependence of model parameter values on climate variables (shaded region in Fig 1B) leads to uncertainty in the boundary of the ecological niche (different shades of orange in Fig 1C).

In our main analyses, we assume that the ecological niche is characterised by its median estimate (black dotted line in Fig 1C). We also validated the derived ecological niche using data from countries with endemic *Ae. aegypti* populations and locations of dengue outbreaks (Fig S7).

### Future climate suitability for *Ae. aegypti*

For each CESM projection and location, we calculated the number of months per year in which climate conditions are projected to be suitable for *Ae. aegypti* survival according to the ecological niche. To check that our model generates results that are consistent with current real-world *Ae. aegypti* observations, we compared the predicted number of months that are suitable for *Ae. aegypti* in different geographical locations globally in 2020 (on average across the CESM simulations) against the locations that are known to have *Ae. aegypti* populations (Fig S8). We then computed the mean change (across the CESM simulations) in the number of months that are projected to be suitable for *Ae. aegypti* survival between 2020 and 2100. We identified the locations that are expected to experience suitable conditions for *Ae. aegypti* for more months of the year in 2100 compared with 2020 (Fig 2 – red). Similarly, we identified the locations that are expected to experience suitable conditions for *Ae. aegypti* for fewer months of the year in 2100 compared with 2020 (Fig 2 – blue). In general, under the assumed ecological model, a poleward expansion of *Ae. aegypti* is expected to occur during the 21^st^ century.

**Figure 2.**
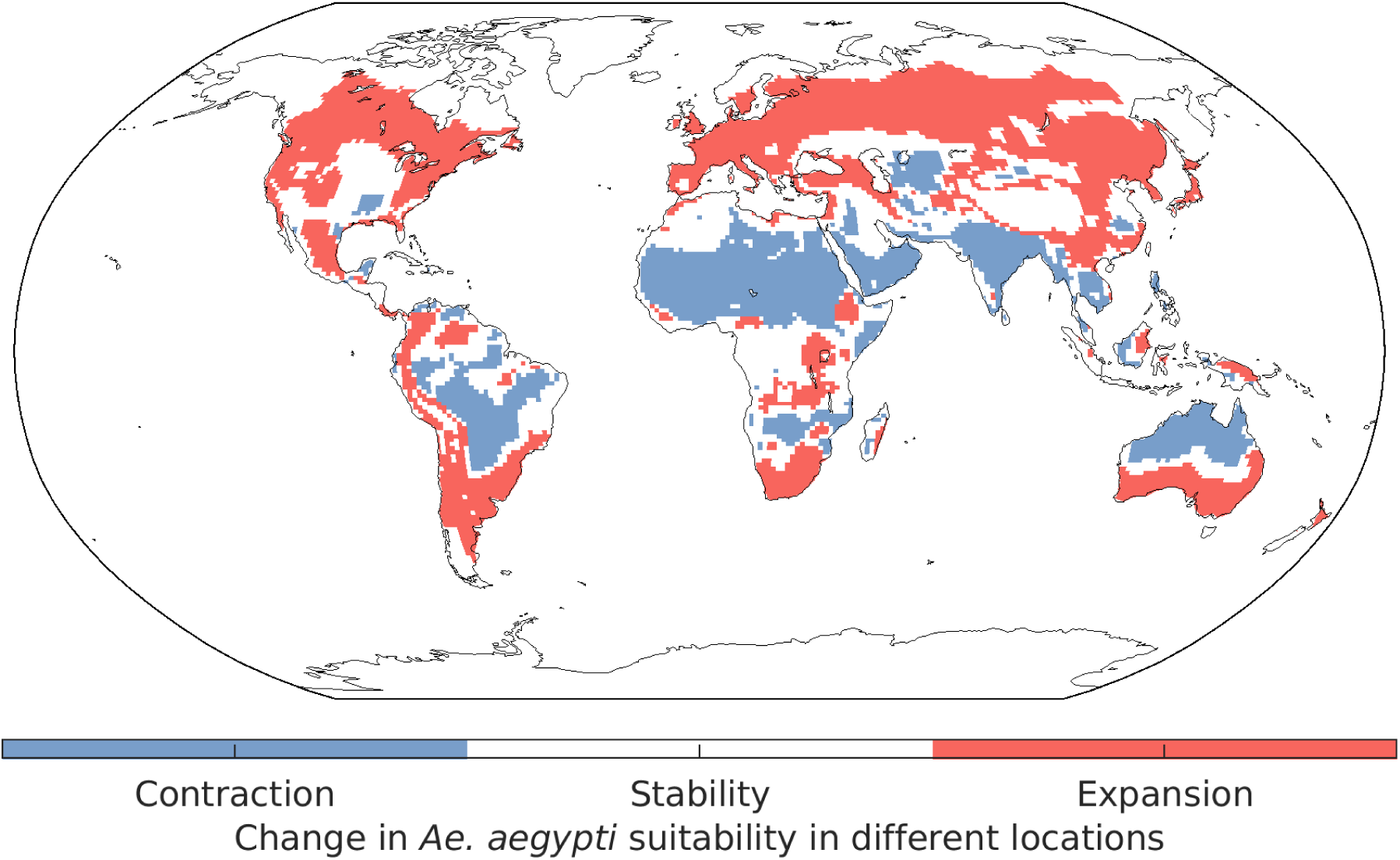
Locations that are expected to see an increase or decrease in the suitability of climatic conditions for *Ae. aegypti*. Locations in which the number of months that are suitable for *Ae. aegypti* increases by more than one in 2100 compared with 2020 are shown in red. Locations with a corresponding decrease are shown in blue. These results were obtained by first calculating the change in the number of suitable months for each CESM projection individually, and then averaging across all projections.

However, future *Ae. aegypti* dynamics depend on natural climate variability, as characterised by the wide range of climate outcomes in the CESM simulations. In each location, we therefore considered the variability in the number of months each year in which climate conditions are projected to be suitable for *Ae. aegypti* survival between CESM projections. In Fig 3A, the maximum number of months (across the CESM projections) that are projected to be suitable for *Ae. aegypti* survival in each location in the year 2100 is shown, with the equivalent minimum number of months depicted in Fig 3B (analogous results for the year 2060 are shown in Fig S9). While Figs 3A and 3B are not representative of any individual CESM projection, they demonstrate the wide variation in projected *Ae. aegypti* suitability in each location between different CESM projections (see also Fig S10).

**Figure 3.**
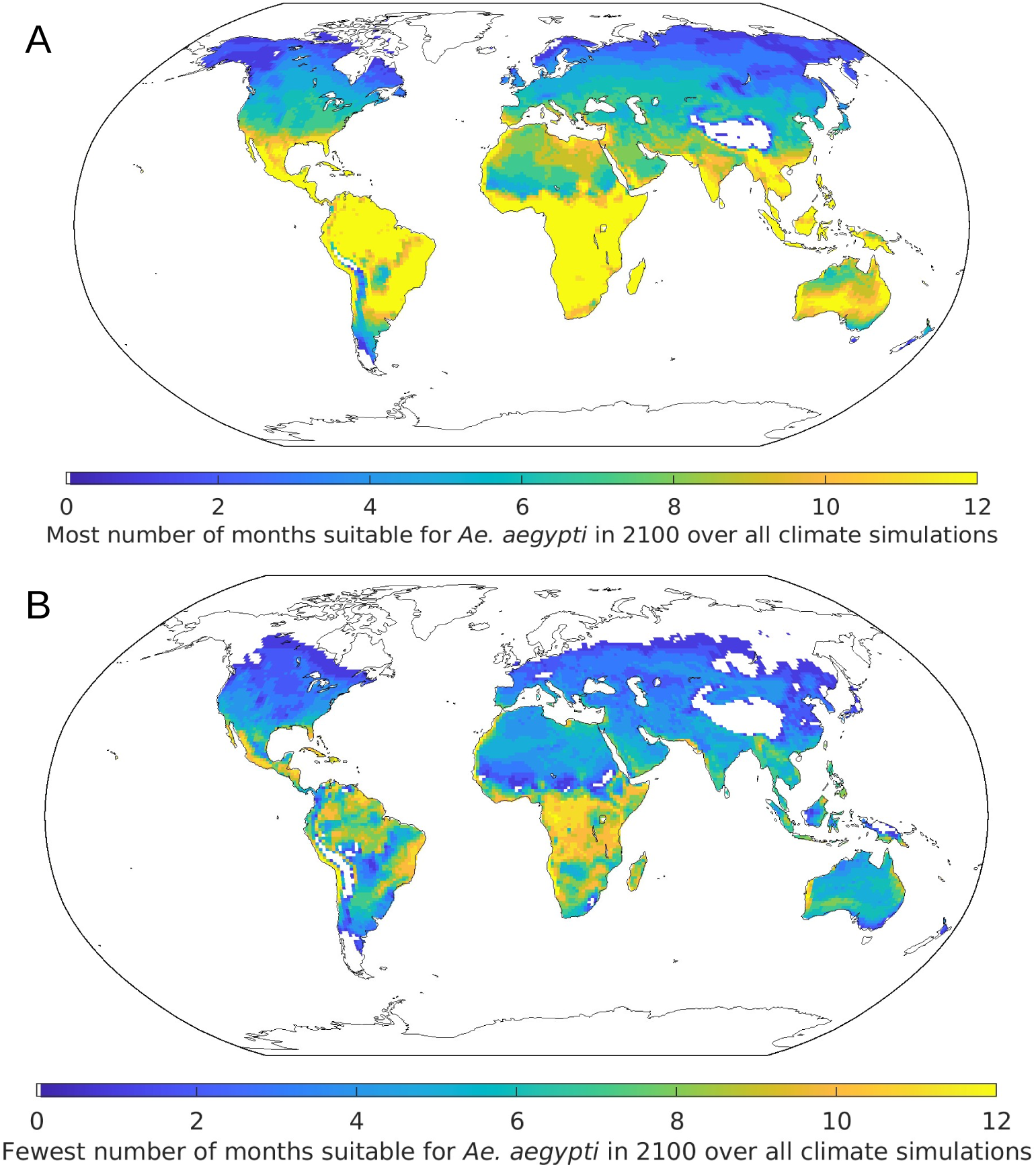
The impact of natural climate variability on future suitability for *Ae. aegypti* in different locations. A. The maximum number of months that are projected to be suitable for *Ae. aegypti* in the year 2100. B. The minimum number of months that are projected to be suitable for *Ae. aegypti* in the year 2100. In both panels, for each latitude-longitude value, the CESM projection corresponding to the most (panel A) or fewest (panel B) number of months that are suitable for *Ae. aegypti* in the year 2100 is chosen.

This highlights the importance of natural climate variability, particularly in geographical locations in which there is a substantial difference between the values shown in Fig 3A and Fig 3B. As an example, in London (in the United Kingdom), where *Ae. aegypti* do not currently inhabit, the number of months suitable for *Ae. aegypti* survival in 2100 could be between one (Fig 3B) and five (Fig 3A) months, depending on natural climate variability. In contrast, if anthropogenic climate change alone is considered (i.e., the mean of the CESM simulations is used to infer future climate suitability for *Ae. aegypti*), then our model projects that four months will be suitable for *Ae. aegypti* in the year 2100. Similarly, in Cape Town (South Africa), the number of months suitable for *Ae. aegypti* survival in 2100 could be between six (Fig 3B) and 11 (Fig 3A) months, compared with eight months based on a projection that does not account for natural climate variability.

## Discussion

Vector-borne pathogens, such as the dengue, Zika and yellow fever viruses, are responsible for over 700,000 deaths each year^22^ and have particularly devastating effects on populations in low- and middle-income countries. Understanding the future threat to planetary public health posed by these pathogens requires changes in the spatial distribution of their vectors to be projected.

In this study, we have constructed a climate-sensitive ecological modelling framework that describes how *Ae. aegypti* populations change as climate variables (temperature and rainfall) vary. We used this model to derive an ecological niche determining the conditions under which self-sustaining *Ae. aegypti* populations are possible (Fig 1C). Using climate projections from the CESM covering the period from 2020-2100, we then projected the locations that will be suitable for *Ae. aegypti* throughout the 21^st^ century and inferred the locations in which suitability for *Ae. aegypti* is expected to change (Fig 2). We also considered the sensitivity of this result to the precise shape of the ecological niche by considering different assumptions about the relationship between rainfall and the rate at which aquatic stage individuals are washed away (Figs S11-S12). In each case that we considered, and similarly to previous modelling studies,^10^ we found that a poleward migration of *Ae. aegypti* is expected.

However, a key difference between our study and previous analyses is that, owing to using a large ensemble of 100 climate projections from the CESM, we were able to conduct a thorough investigation into the impact of natural climate variability on projections of the global spatial distribution of *Ae. aegypti*. Natural climate variability arises due to internal atmospheric processes and interactions between different components of the climate system (i.e., atmospheric, oceanic, land and cryospheric processes and their coupled interactions). It is well-studied by climate scientists and is known to be as important as anthropogenic climate change in shaping Earth’s future climate trajectory, especially regionally and even over many decades.^15, 23^ Only one previous study has considered natural climate variability in the context of future *Ae. aegypti* dynamics,^24^ but only a small number of climate model projections were available for use in that research, meaning that the full effect of natural climate variability on future environmental suitability for *Ae. aegypti* had not been properly assessed.

We found that natural climate variability has a substantial effect on the suitability of different locations to harbour *Ae. aegypti* in the future (Fig 3). When estimates are constructed accounting for anthropogenic climate change but neglecting natural climate variability, the wide variation in possible climatic conditions in different locations in future years is ignored. This is important, particularly as some locations may be more suitable for *Ae. aegypti* in future due to the combination of anthropogenic climate change and natural climate variability. As a result, outbreaks of vector-borne disease may occur in some places earlier than expected under anthropogenic climate change alone.

Previous research has involved projecting the locations of pathogen vectors under a changing climate (without natural climate variability) and considering the impacts of climate change on epidemiological parameters. For example, Kraemer *et al.*^25^ developed a machine learning model to predict the current spatial distributions of both *Ae. aegypti* and *Ae. albopictus* and then used it to project future changes in those distributions accounting for climate change, urbanisation and increased connectivity between locations.^13^ Mordecai *et al.*^21^ and Ryan *et al.*^10^ considered models in which the basic reproduction number (𝑅_!_) of *Aedes*-borne viruses is assumed to depend on temperature, and projected the future temperature suitability of locations in the Americas and globally for pathogen transmission. Parham and Michael^26^ developed a similar model for malaria in which 𝑅_!_ depends on temperature and rainfall, but did not use it to make global predictions. Other research includes the development of the metric Index P, which has been used to estimate the suitability of climate conditions for transmission of pathogens including the dengue^27, 28^ and West Nile^29, 30^ viruses. While these and other similar studies have provided valuable insights into the effects of climate change on vector ecology and the potential for disease outbreaks, they did not consider natural climate variability.

Like any quantitative analysis, our modelling study involved some assumptions and simplifications. Our main goal was to investigate whether natural climate variability affects projections of future vector dynamics rather than to make precise quantitative predictions. Consequently, additional details should be included in the model if it is used to guide vector or pathogen surveillance programmes. For example, the presence of *Ae. aegypti* does not only depend on climate suitability, but also on a range of factors including human behaviour.

Future iterations of our modelling framework could account for changing human population sizes, geographical heterogeneity in socioeconomic development^14, 31^ and/or methods of water storage in different locations (water storage containers act as breeding sites for *Ae. aegypti*).^32^ We chose to study *Ae. aegypti* here as it is the primary vector of a range of pathogens.^33^ However, pathogens can be spread by multiple vectors, and so our framework could be extended to other *Aedes* species (such as *Ae. albopictus*, which transmits pathogens and are known to be particularly invasive) or non-*Aedes* vectors.

While our model projections indicate that *Ae. aegypti* are likely to spread to new locations, we also note that climate conditions in some places are projected to become less suitable for *Ae. aegypti* during the 21^st^ century (Fig 2). We urge against interpretating this result to mean that the situation will become “better” in those locations. First, our model did not include the possibility that the vectors might adapt to allow survival in these adverse climate conditions,^34^ which may be particularly relevant over the long timescale that we are considering. Second, we note that if climate conditions become unsuitable for *Ae. aegypti*, for example due to temperatures becoming too hot, then a range of other problems will arise, including increased drought, storm severity, food insecurity and population displacement.

Despite these simplifications, we have demonstrated that the future spatial distribution of vectors of globally important pathogens depends not only on anthropogenic climate change but on the combined effects of climate change and natural climate variability. It is important that future projections of climate-sensitive ecological and epidemiological systems consider natural climate variability. During the 21^st^ century, vectors and the pathogens that they transmit will undoubtedly spread to new locations due to climate shifts. Careful climate monitoring and rigorous surveillance for vectors and vector-borne pathogens is essential.

Surveillance strategies should be informed by the wide range of possible future climate scenarios, accounting for all potential sources of variability. This will allow more resilient public health infrastructure to be built than considering a single possible climate future alone. This is of clear importance for planetary public health.

## AUTHORS’ CONTRIBUTIONS

ARK – formal analysis, investigation, software, validation, writing – original draft, writing – review and editing.

UO – methodology, writing – review and editing.

LS – methodology, investigation, writing – review and editing.

JWH – conceptualization, methodology, writing – review and editing.

MJT – supervision, writing – review and editing.

RNT – conceptualization, methodology, project administration, supervision, writing – original draft, writing – review and editing.

## ACKNOWLEDGEMENTS

Thanks to members of the Zeeman Institute for Systems Biology and Infectious Disease Epidemiology Research at the University of Warwick, and the Wolfson Centre for Mathematical Biology at the University of Oxford, for useful discussions about this research.

## DATA SHARING

All data generated or analysed during this study, including computing code for reproducing our results, are available at: www.github.com/KayeARK/Climate_Change_NCV_Ae_Aegypti

## References

1 Ryan SJ, Lippi CA, Caplan T, et al. The current landscape of software tools for the climate-sensitive infectious disease modelling community. Lancet Planet Health 2023; 7: e527–36.

2 Mordecai EA, Caldwell JM, Grossman MK, et al. Thermal biology of mosquito-borne disease. Ecol Lett 2019; 22: 1690–708.

3 Yang HM, Macoris MLG, Galvani KC, Andrighetti MTM, Wanderley DMV. Assessing the effects of temperature on the population of *Aedes aegypti*, the vector of dengue. Epidemiol Infect 2009; 137: 1188–202.

4 Tesla B, Demakovsky LR, Mordecai EA, et al. Temperature drives Zika virus transmission: Evidence from empirical and mathematical models. Proc R Soc B Biol Sci 2018; 285: p.

5 Juliano SA, O’Meara GF, Morrill JR, Cutwa MM. Desiccation and thermal tolerance of eggs and the coexistence of competing mosquitoes. Oecologia 2002; 130: 458–69.

6 Alto BW, Juliano SA. Precipitation and temperature effects on populations of *Aedes albopictus* (Diptera: Culicidae): implications for range expansion. J Med Entomol 2001; 38: 646–56.

7 Gouagna LC, Dehecq JS, Fontenille D, Dumont Y, Boyer S. Seasonal variation in size estimates of *Aedes albopictus* population based on standard mark-release-recapture experiments in an urban area on Reunion Island. Acta Trop 2015; 143: 89–96.

8 Guerra CA, Reiner RC, Perkins TA, et al. A global assembly of adult female mosquito mark-release-recapture data to inform the control of mosquito-borne pathogens. Parasit Vectors 2014; 7: 276–276.

9 Yang B, Borgert BA, Alto BW, et al. Modelling distributions of *Aedes aegypti* and *Aedes albopictus* using climate, host density and interspecies competition. PLoS Negl Trop Dis 2021; 15: e0009063.

10 Ryan SJ, Carlson CJ, Mordecai EA, Johnson LR. Global expansion and redistribution of Aedes-borne virus transmission risk with climate change. PLoS Negl Trop Dis 2018; 13: e0007213.

11 Brady OJ, Johansson MA, Guerra CA, et al. Modelling adult *Aedes aegypti* and *Aedes albopictus* survival at different temperatures in laboratory and field settings. Parasit Vectors 2013; 6: 351–351.

12 Mordecai EA, Paaijmans KP, Johnson LR, et al. Optimal temperature for malaria transmission is dramatically lower than previously predicted. Ecol Lett 2013; 16: 22–30.

13 Kraemer MUG, Reiner RC, Brady OJ, et al. Past and future spread of the arbovirus vectors *Aedes aegypti* and *Aedes albopictus*. Nat Microbiol 2019; 4: 854–854.

14 Liu-Helmersson J, Brännström Å, Sewe MO, Semenza JC, Rocklöv J. Estimating past, present, and future trends in the global distribution and abundance of the arbovirus vector *Aedes aegypti* under climate change scenarios. Front Public Health 2019; 7: 148.

15 Hawkins E, Sutton R. The potential to narrow uncertainty in regional climate predictions. Bull Am Meteorol Soc 2009; 90: 1095–108.

16 Deser C, Hurrell JW, Phillips AS. The role of the North Atlantic Oscillation in European climate projections. Clim Dyn 2017; 49: 3141–57.

17 Ricke KL, Caldeira K. Natural climate variability and future climate policy. Nat Clim Change 2014; 4: 333–8.

18 Hurrell JW, Holland MM, Gent PR, et al. The Community Earth System Model: A framework for collaborative research. Bull Am Meteorol Soc 2013; 94: 1339–60.

19 Danabasoglu G, Lamarque J -F., Bacmeister J, et al. The Community Earth System Model Version 2 (CESM2). J Adv Model Earth Syst 2020; 12: e2019MS001916.

20 Rodgers KB, Lee S-S, Rosenbloom N, et al. Ubiquity of human-induced changes in climate variability. Earth Syst Dyn 2021; 12: 1393–411.

21 Mordecai EA, Cohen JM, Evans MV, et al. Detecting the impact of temperature on transmission of Zika, dengue, and chikungunya using mechanistic models. PLoS Negl Trop Dis 2017; 27: e0005568.

22 World Health Organization. Fact sheet: Vector-borne diseases. 2020 www.who.int/news-room/fact-sheets/detail/vector-borne-diseases.

23 Rondeau-Genesse G, Braun M. Impact of internal variability on climate change for the upcoming decades: analysis of the CanESM2-LE and CESM-LE large ensembles. Clim Change 2019; 156: 299–314.

24 Monaghan AJ, Sampson KM, Steinhoff DF, et al. The potential impacts of 21st century climatic and population changes on human exposure to the virus vector mosquito *Aedes aegypti*. Clim Change 2018; 146: 487–500.

25 Kraemer MUG, Sinka ME, Duda KA, et al. The global distribution of the arbovirus vectors *Aedes aegypti* and *Ae. albopictus*. eLife 2015; 4: e08347–e08347.

26 Parham PE, Michael E. Modeling the effects of weather and climate change on malaria transmission. Environ Health Perspect 2010; 118: 620–6.

27 Obolski U, Perez PN, Villabona-Arenas CJ, Thézé J, Faria NR, Lourenço J. MVSE: An R- package that estimates a climate-driven mosquito-borne viral suitability index. Methods Ecol Evol 2019; 10: 1357–70.

28 Nakase T, Giovanetti M, Obolski U, Lourenço J. Global transmission suitability maps for dengue virus transmitted by *Aedes aegypti* from 1981 to 2019. Sci Data 2023; 10: 275.

29 Lourenço J, Thompson RN, Thézé J, Obolski U. Characterising West Nile virus epidemiology in Israel using a transmission suitability index. Eurosurveillance 2020; 25: 1900629.

30 Lourenço J, Barros SC, Zé-Zé L, et al. West Nile virus transmission potential in Portugal. Commun Biol 2022; 5: 6.

31 Messina JP, Brady OJ, Golding N, et al. The current and future global distribution and population at risk of dengue. Nat Microbiol 2019; 4: 1508–15.

32 Koenraadt CJM, Tuiten W, Sithiprasasna R, Kijchalao U, Jones JW, Scott TW. Dengue knowledge and practices and their impact on *Aedes aegypti* populations in Kamphaeng Phet, Thailand. Am J Trop Med Hyg 2006; 74: 692–700.

33 Souza-Neto JA, Powell JR, Bonizzoni M. *Aedes aegypti* vector competence studies: A review. Infect Genet Evol 2019; 67: 191–209.

34 Couper LI, Farner JE, Caldwell JM, et al. How will mosquitoes adapt to climate warming? eLife 2021; 10: e69630.

