## Supplementary Material for "The impact of climate change and natural climate variability on the global distribution of *Aedes aegypti*"

[3]Department of Epidemiology and Preventive Medicine, School of Public Health, Faculty of Medicine, Tel Aviv University, Tel Aviv-Yafo, 6997801, Israel

[4]Porter School of the Environment and Earth Sciences, Faculty of Exact Sciences, Tel Aviv University, Tel Aviv-Yafo, 6997801, Israel

### Supplementary Text

#### Supplementary details about the ecological model and its parameterisation

##### The ecological model

As described in the main text, in the ecological model the *Ae. aegypti* population is divided according to their life cycle stage: eggs ($E$), aquatic stage (larvae or pupae; $A$) and adult female mosquitoes ($M$). The compartmental, ordinary differential equation model is given by

$$\frac{dE}{dt} = a(T)M - b(T)E,$$

$$\frac{dA}{dt} = b(T)E\left(1 - \frac{A}{K(R)}\right) - c(R)A - d(T)A - f(T)A,$$

$$\frac{dM}{dt} = \frac{1}{2}f(T)A - g(T)M,$$

in which $T$ indicates dependence on temperature (°C) and $R$ indicates dependence on rainfall (mm day$^{-1}$).

The parameter $a(T)$ is the birth rate (eggs per adult female per day), $b(T)$ is the egg-to-aquatic development rate (in a setting in which the birth rate is not resource limited), $d(T)$ is the aquatic stage death rate, $f(T)$ is the aquatic-to-adult development rate and $g(T)$ is the adult death rate. Rainfall-dependence is included through the aquatic stage carrying capacity ($K(R)$) and the rate at which aquatic stage individuals are washed away ($c(R)$). The factor of $\frac{1}{2}$ is included in the final equation as we are predominantly interested in female adults; male adults do not spread pathogens. However, we include male *Ae. aegypti* in the model up to the adult stage, since they contribute to competition for resources.

For fixed temperature and rainfall values, the non-zero equilibrium value of $M$ under this model is given by

$$M^* = K(R)\left(\frac{f(T)}{2g(T)} - \frac{c(R) + d(T) + f(T)}{a(T)}\right).$$

The value of $M^*$ is positive, so that a self-sustaining *Ae. aegypti* population is possible, when

$$\frac{f(T)}{2g(T)} > \frac{c(R) + d(T) + f(T)}{a(T)}.$$

This expression is used as the basis for the ecological niche shown in Fig 1C in the main text.

Temperature-dependent model parameters

The relationships between temperature and each temperature-dependent model parameter are determined based on the data reported by Mordecai *et al.*[1] Details for each model parameter are given below (further information about the functional forms of the fitted relationships are provided in the following subsection):

- Birth rate, $a(T)$: We fitted a Brière equation directly to data describing the number of eggs laid per adult female per day at different temperature values.
- Egg-to-aquatic development rate, $b(T)$: Data describing the egg-to-aquatic development rate were not available from the Mordecai *et al.*[1] study. We therefore estimated $b(T)$ indirectly by first fitting a Brière equation to data describing the egg-to-adult development rate as a function of temperature ($\gamma(T)$, say). Neglecting resource limitation, and the possibility that aquatic stage individuals die or are washed away, the egg-to-adult development rate depends on both the egg-to-aquatic and aquatic-to-adult development rates. A previous study by Silva

*et al.*[2] in which climate dependence was not considered found that mean egg-to-aquatic and aquatic-to-adult development rates were 0.24 day$^{-1}$ and 0.125 day$^{-1}$, respectively. Assuming that the ratio between these quantities applies at all temperatures, then $b(T) = \frac{0.125+0.24}{0.125}\gamma(T) = \frac{73}{25}\gamma(T)$.

- Aquatic-to-adult development rate, $f(T)$: Analogously to estimating the egg-to-aquatic development rate, $f(T) = \frac{73}{48}\gamma(T)$.
- Aquatic stage death rate, $d(T)$. Data were available[1] describing the probability that an individual survives from egg to adult at different temperatures ($p(T)$, say). We fitted a quadratic equation to these data. Neglecting resource limitation, and the possibility that aquatic stage individuals are washed away, gives $p(T) = \frac{f(T)}{d(T)+f(T)}$ and so $d(T) = f(T)\left(\frac{1}{p(T)} - 1\right)$.
- Adult death rate, $g(T)$. A quadratic equation was fitted directly to data describing the adult lifespan ($1/g(T)$) at different temperature values.

Parameters fits are shown for $a(T)$ (Fig S2), $\gamma(T)$ (Fig S3), $p(T)$ (Fig S4) and $1/g(T)$ (Fig S5), from which the posteriors for the temperature-dependent parameters of the ecological model were obtained (Fig S6A-D).

Details of the temperature-dependent parameter fitting

As described above, a Brière equation or a quadratic equation was fitted to determine the relationship between individual *Ae. aegypti* model parameters and temperature. Both of these functions have a similar peaked shape, but Brière equations are asymmetric whereas quadratic equations are symmetric about the peak value. We therefore chose whether to fit a Brière or quadratic equation to determine the relationship between temperature and each fitted parameter according to whether the relevant data appeared to be symmetric about their peak. This approach was also used by Mordecai *et al.*[1]

The functional form of the fitted Brière equation for the *Ae. aegypti* birth rate is

$$a(T) = \max\left(0, \mathrm{Re}\left(\alpha T(T - T_0)\sqrt{T_m - T}\right) + \mathcal{N}(0, \sigma^2)\right), \qquad \text{(S1)}$$

in which $\mathrm{Re}(z)$ denotes the real part of the complex number $z$, and $\mathcal{N}(0, \sigma^2)$ represents Gaussian noise with mean zero and variance $\sigma^2$. An analogous equation was used when we characterised $\gamma(T)$ using a Brière equation. Similarly, the quadratic functional form for $p(T)$ is

$$p(T) = \max\left(0, -\alpha(T - T_0)(T - T_m) + \mathcal{N}(0, \sigma^2)\right), \qquad \text{(S2)}$$

in which $T_m > T_0$, with a similar equation for $1/g(T)$.

Here, we describe the details of the MCMC approach used to determine the sub-parameters $\alpha$, $T_0$, $T_m$ and $\sigma^2$ of $a(T)$. A similar method was used for all fitted temperature-dependent responses (and we reused the same notation for the sub-parameters – i.e., $\alpha$, $T_0$, $T_m$ and $\sigma^2$ – in each case).

The Metropolis-Hastings algorithm was used. In each step of the MCMC chain, new values of $\alpha$, $T_0$, $T_m$ and $\sigma^2$ were proposed, each sampled from independent Gaussian proposal distributions with mean equal to the current value and variances $5 \times 10^{-3}$, 1, 0.1 and 1, respectively. These variance values (referred to as Σ values in Tables S1 and S2) were chosen to achieve an acceptance rate of around 0.234.[3] The Σ values for other fitted temperature-dependent parameters are shown in Tables S1 and S2, along with the priors used for each sub-parameter. The algorithm was run for 100,000 steps, including a burn-in of 50,000 steps.

We repeated the fitting procedure a further four times, each time starting the MCMC chain from a different initial state, allowing us to confirm convergence of the original chain using the Gelman-Rubin statistic (see captions to Figs S2-S5). When fitting the temperature-dependent response for $a(T)$, the initial value of $\alpha$ was sampled for each chain from a $U(1 \times 10^{-3}, 1 \times 10^{-1})$ distribution, $T_0$ was sampled from a $U(0,20)$ distribution, $T_m$ was sampled from a $U(20,40)$ distribution and $\sigma^2$ was sampled from a $U(1,10)$ distribution. The distributions from which the initial values were sampled for the other temperature-dependent responses are shown in Tables S1 and S2.

| Parameter | Sub-parameter | Prior | Initial value | Σ Value |
|---|---|---|---|---|
| Birth rate, $a(T)$ | $\alpha$ | $\Gamma(2, 10^{-2})$ | $\mathrm{U}(10^{-3}, 10^{-1})$ | $5 \times 10^{-3}$ |
| | $T_0$ | $\Gamma(10, 2)$ | $\mathrm{U}(0, 20)$ | 1 |
| | $T_m$ | $\Gamma(10, 4)$ | $\mathrm{U}(20, 40)$ | 0.1 |
| | $\sigma^2$ | $\mathrm{U}(0, 10)$ | $\mathrm{U}(1, 10)$ | 1 |
| Egg-to-adult development rate, $\gamma(T)$ | $\alpha$ | $\Gamma(9, 10^{-5})$ | $\mathrm{U}(10^{-5}, 10^{-3})$ | $5 \times 10^{-6}$ |
| | $T_0$ | $\Gamma(7, 2)$ | $\mathrm{U}(0, 20)$ | 0.5 |
| | $T_m$ | $\Gamma(10, 4)$ | $\mathrm{U}(20, 50)$ | 0.5 |
| | $\sigma^2$ | $\mathrm{U}(0, 1)$ | $\mathrm{U}(0, 1)$ | 0.01 |

**Table S1. Technical details of the MCMC procedure used to determine the temperature-dependent responses of $\boldsymbol{a(T)}$ and $\boldsymbol{\gamma(T)}$.** Due to the asymmetric nature of the data for these parameters, Brière equations were used (equation S1). The notation $\Gamma(x, y)$ represents a gamma distribution with shape parameter $x$ and scale parameter $y$, and $\mathrm{U}(x, y)$ is a continuous uniform distribution with bounds $x$ and $y$. The initial value in the MCMC chain of each sub-parameter was sampled from the distributions listed in the fourth column.

| Parameter | Sub-parameter | Prior | Initial value | Σ Value |
|---|---|---|---|---|
| Egg-to-adult survival probability, $p(T)$ | $\alpha$ | $\Gamma(7, 0.001)$ | $\mathrm{U}(0.0001, 0.01)$ | 0.0001 |
| | $T_0$ | $\Gamma(7, 2)$ | $\mathrm{U}(0, 20)$ | 0.5 |
| | $T_m$ | $\Gamma(10, 4)$ | $\mathrm{U}(20, 50)$ | 0.5 |
| | $\sigma^2$ | $\mathrm{U}(0, 5)$ | $\mathrm{U}(0, 2)$ | 0.01 |
| Adult lifespan, $1/g(T)$ | $\alpha$ | $\Gamma(1, 0.5)$ | $\mathrm{U}(0.01, 1)$ | 0.015 |
| | $T_0$ | $\Gamma(5, 2)$ | $\mathrm{U}(0, 20)$ | 1 |
| | $T_m$ | $\Gamma(9, 5)$ | $\mathrm{U}(20, 50)$ | 1 |
| | $\sigma^2$ | $\mathrm{U}(0, 50)$ | $\mathrm{U}(0, 20)$ | 1 |

**Table S2. Technical details of the MCMC procedure used to determine the temperature-dependent responses of $\boldsymbol{p(T)}$ and $\boldsymbol{1/g(T)}$.** Due to the symmetric nature of the data for these parameters, quadratic equations were used (equation S2). The notation $\Gamma(x, y)$ represents a gamma distribution with shape parameter $x$ and scale parameter $y$, and $\mathrm{U}(x, y)$ is a continuous uniform distribution with bounds $x$ and $y$. The initial value in the MCMC chain of each sub-parameter was sampled from the distributions listed in the fourth column.

#### Rainfall-dependent model parameters

Two ecological model parameters were assumed to depend on rainfall: the aquatic stage carrying capacity ($K(R)$) and the rate at which aquatic stage individuals are washed away (the larval flush out rate, $c(R)$). Relationships between the amount of rainfall and the values of these parameters were derived using the approach of Tompkins and Ermert.[4]

##### *Aquatic stage carrying capacity, $K(R)$*

The aquatic stage carrying capacity is defined as $K(R) = k(R)D$, in which $k(R)$ is the carrying capacity per unit area and $D$ is the area of the location under consideration. We defined $k(R) = w(R)\frac{M_L}{m}$, with $w(R)$ denoting the proportion of land covered with *Ae. aegypti* breeding sites, $M_L$ denoting the total mass of aquatic stage individuals that can be present at a breeding site and $m$ denoting the average mass of an individual in the aquatic phase. Following Tompkins and Ermert,[4]

$$\frac{\mathrm{d}w(R)}{\mathrm{d}t} = \kappa\left[R\left(w_{max} - w(R)\right) - w(R)(\eta + \zeta)\right],$$

in which $w_{max}$ is the proportion of the land surface that is covered by depressions that can become filled with water. The parameter $\kappa$ is based on the geometry of the depressions, and sets the overall rate at which they are filled with or lose water, and the parameters $\eta$ and $\zeta$ set the relative rates at which water evaporates and is infiltrated into the ground, respectively.

The equilibrium value of $w(R)$ is then

$$w(R) = \frac{Rw_{max}}{\eta + \zeta + R},$$

so that the aquatic stage carrying capacity per unit area is

$$k(R) = \frac{Rw_{max}}{\eta + \zeta + R}\frac{M_L}{m},$$

and

$$K(R) = \frac{Rw_{max}}{\eta + \zeta + R}\frac{M_L D}{m}.$$

We set $\eta = 5$ mm day$^{-1}$,[4] $\zeta = 245$ mm day$^{-1}$,[4] $M_L = 300$ mg m$^{-2}$,[4] and $m = 4.59$ mg (this is the average mass from a sample of 1000 male and 1000 female pupae[5]). We set $w_{max} = \frac{1}{25}$, so that 4% of the land's surface can become filled with water.[4]

In the expression above, in the absence of rainfall, then $K(0) = 0$. When there is a very large amount of rainfall, $K(R)$ tends to a constant value (the maximum possible carrying capacity). The dependence of $K(R)$ on $R$ is shown for values of $R$ between 0 and 25 mm day$^{-1}$ in Fig S6E. For comparison, we also plot the (constant) carrying capacity from a model developed by Silva *et al.*[2] The study location in that research was Nova Iguaçu, Brazil which has an average daily rainfall of 4.89mm. This is consistent with the corresponding value of $K(R)$ in our model (Fig S6E).

*Larval flush out rate,* $c(R)$

Each day, the probability that an aquatic stage individual survives being washed away is assumed to have the form[4]

$$K_f(R) = C_1 + C_2 \exp(-C_3 R).$$

We assume that, in the absence of rainfall, individuals will not be washed away, and for large amounts of rainfall, individuals will definitely be washed away. Consequently, $K_f(0) = 1$ and $K_f(R) \to 0$ as $R \to \infty$. Setting $C_3 = 1$, so that the daily survival probability is 0.25 at a moderate rainfall level of 500 mm year$^{-1}$,[6] gives $K_f(R) = \exp(-R)$.

We note that, in the ecological model, larval flush out occurs at (exponential) rate $c(R)$. Hence, the probability of an aquatic stage individual surviving any single day is $\exp(-c(R))$. Matching this with the expression above gives $c(R) = R$ (Fig S6F).

We also conducted supplementary analyses in which we considered the sensitivity of our results to the shape of the ecological niche by considering different assumptions about the relationship between rainfall and the rate at which aquatic stage individuals are washed away. Specifically, we also considered scenarios in which $c(R) = 2R$ (Figs S11A and S12A) and $c(R) = \frac{1}{2}R$ (Figs S11B and S12B). In each case that we considered, our qualitative findings about the likely future poleward spread of *Ae. aegypti* and the substantial impact of natural climate variability were unchanged.

#### **Supplementary Figures**

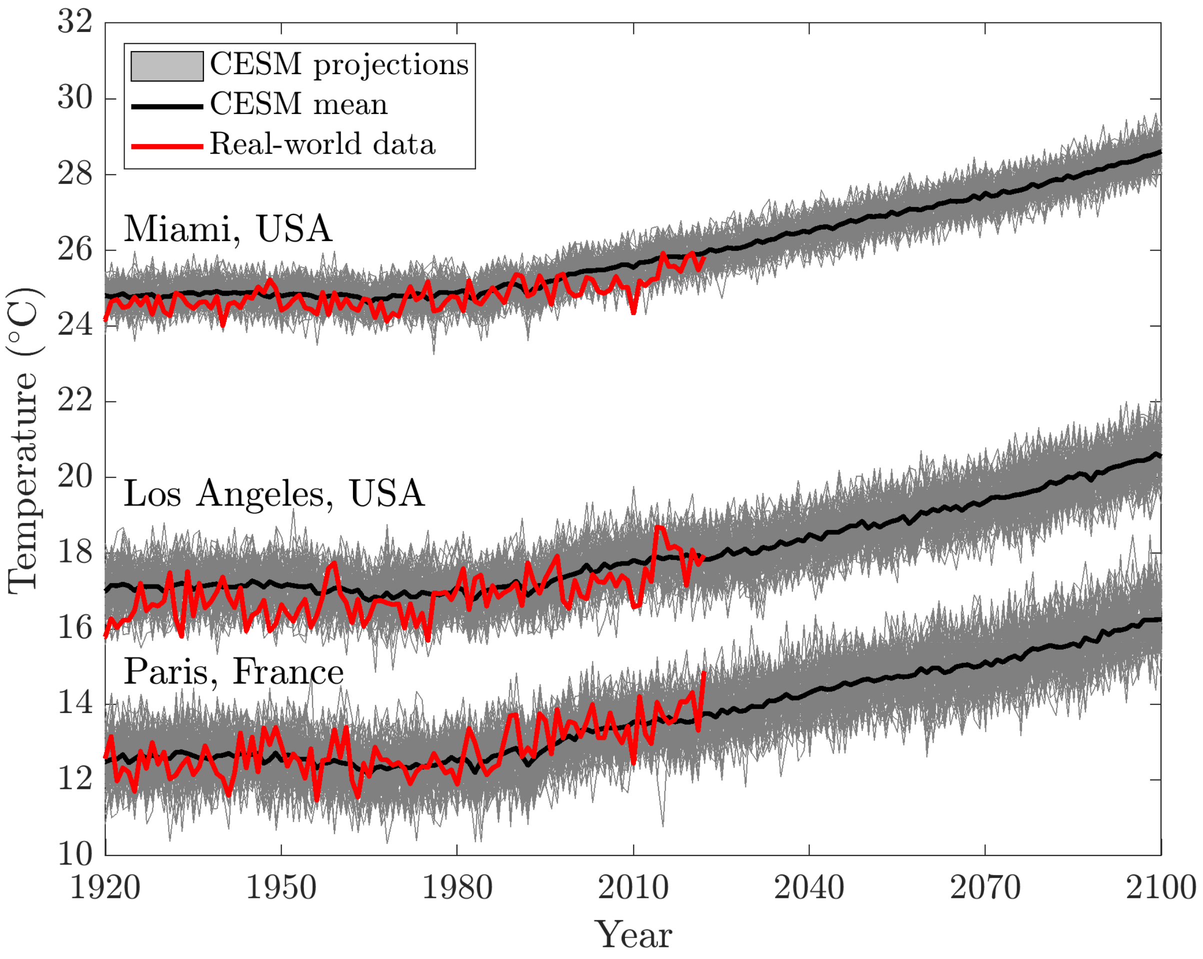


**Figure S1. The variability in real-world climate data exceeds that of the CESM ensemble mean.** As an example, temperature data were extracted from the CESM LENS2 dataset in the locations of Miami, Los Angeles and Paris covering the period from 1920-2100, and the yearly mean was considered. This figure indicates the signal due to anthropogenic climate change (mean of the CESM projections – black), the individual CESM projections (grey) and the temperature observed in the real-world from 1920-2022 (extracted from the Berkeley Earth Surface Temperatures online database;[7] red). This indicates that the variability in real-world climate data is more accurately represented by the variability across the CESM simulations, rather than the ensemble mean.

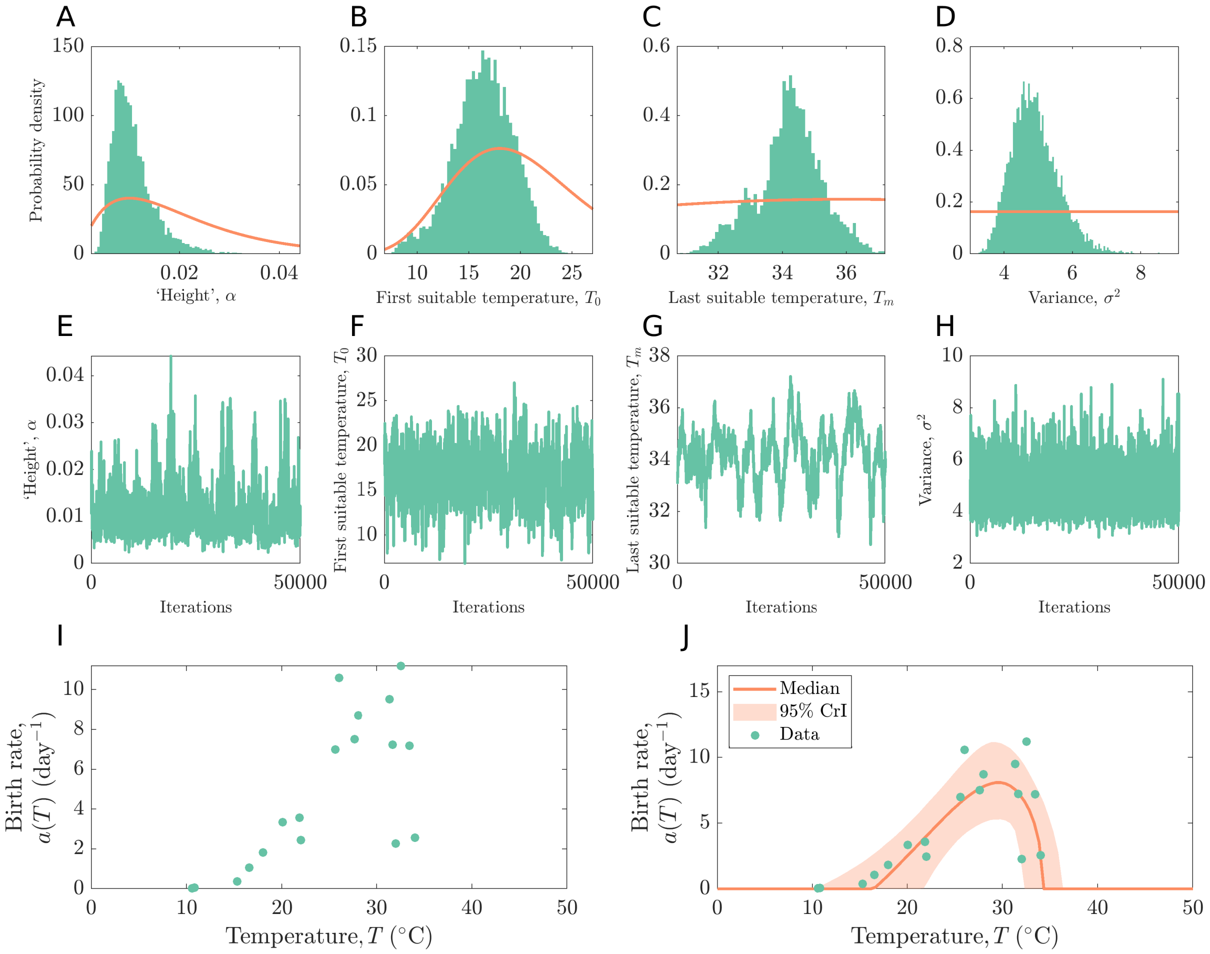


**Figure S2. The dependence of the *Ae. aegypti* birth rate ($a(T)$) on temperature.** A-D. Prior (red) and posterior (green) distributions for each of the fitted sub-parameters ($\alpha$, $T_0$, $T_m$ and $\sigma^2$). To allow the posterior distribution to be seen clearly, x-axes limits are restricted to the minimum and maximum values in the posterior. E-H. Trace plots corresponding to the posterior distributions shown in panels A-D. 100,000 steps were run in the MCMC chain, with the first 50,000 discarded as burn-in (acceptance rate: 0.2419). Five chains were run to compute the Gelman-Rubin statistic (which was 1.0141, 1.0015, 1.0222 and 1.0034 for $\alpha$, $T_0$, $T_m$ and $\sigma^2$, respectively); the trace plots in panels E-H are from the first chain. I. Data describing the *Ae. aegypti* birth rate as a function of temperature. J. Brière equation fit to the data in panel I (data – green; median fit – red; 95% equal-tailed credible interval – shaded region).

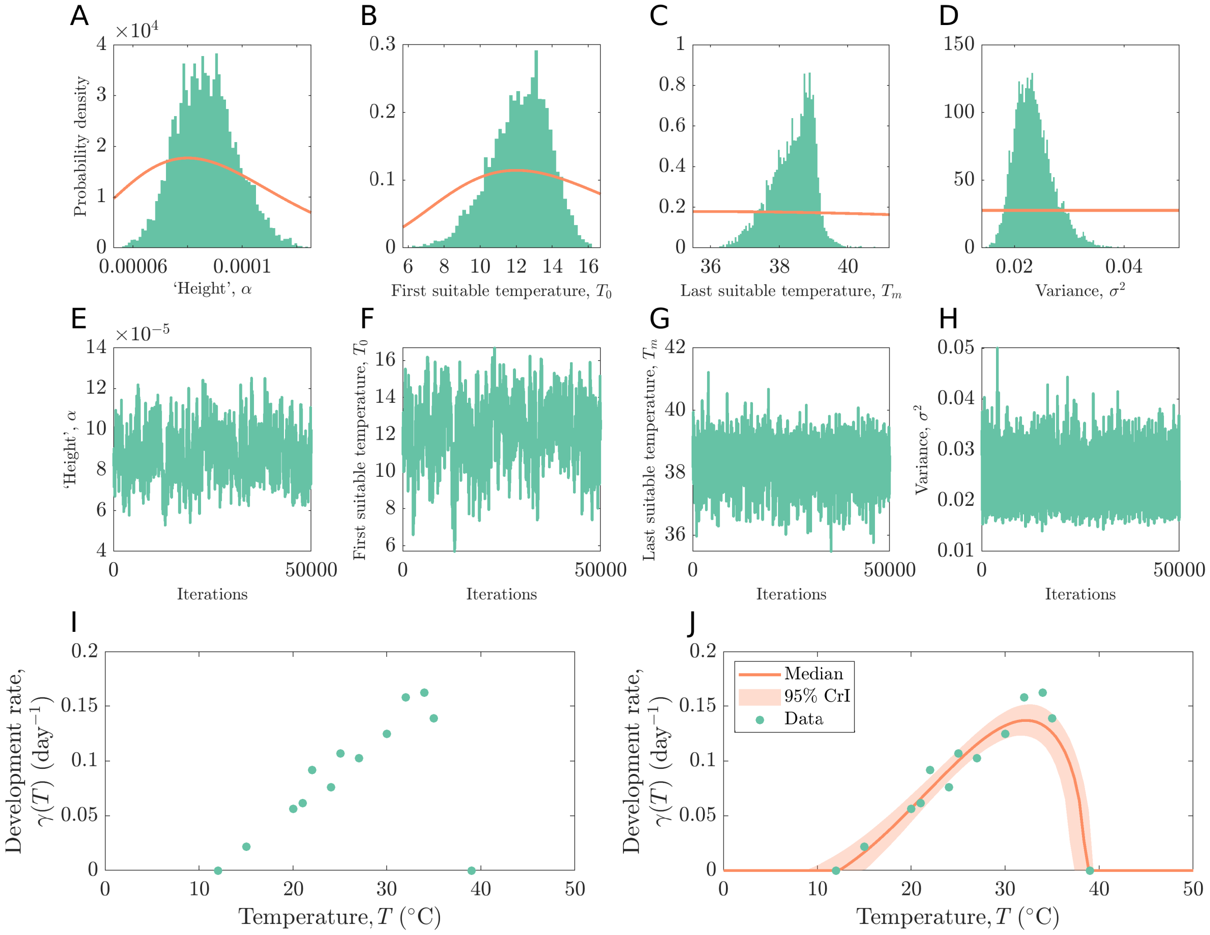


**Figure S3. The dependence of the *Ae. aegypti* egg-to-adult development rate ($\gamma(T)$) on temperature.** A-D. Prior (red) and posterior (green) distributions for each of the fitted sub-parameters ($\alpha$, $T_0$, $T_m$ and $\sigma^2$). To allow the posterior distribution to be seen clearly, x-axes limits are restricted to the minimum and maximum values in the posterior. E-H. Trace plots corresponding to the posterior distributions shown in panels A-D. 100,000 steps were run in the MCMC chain, with the first 50,000 discarded as burn-in (acceptance rate: 0.1982). Five chains were run to compute the Gelman-Rubin statistic (which was 1.0008, 1.0004, 1.0010 and 1.0002 for $\alpha$, $T_0$, $T_m$ and $\sigma^2$, respectively); the trace plots in panels E-H are from the first chain. I. Data describing the *Ae. aegypti* egg-to-adult development rate as a function of temperature. J. Brière equation fit to the data in panel I (data – green; median fit – red; 95% equal-tailed credible interval – shaded region).

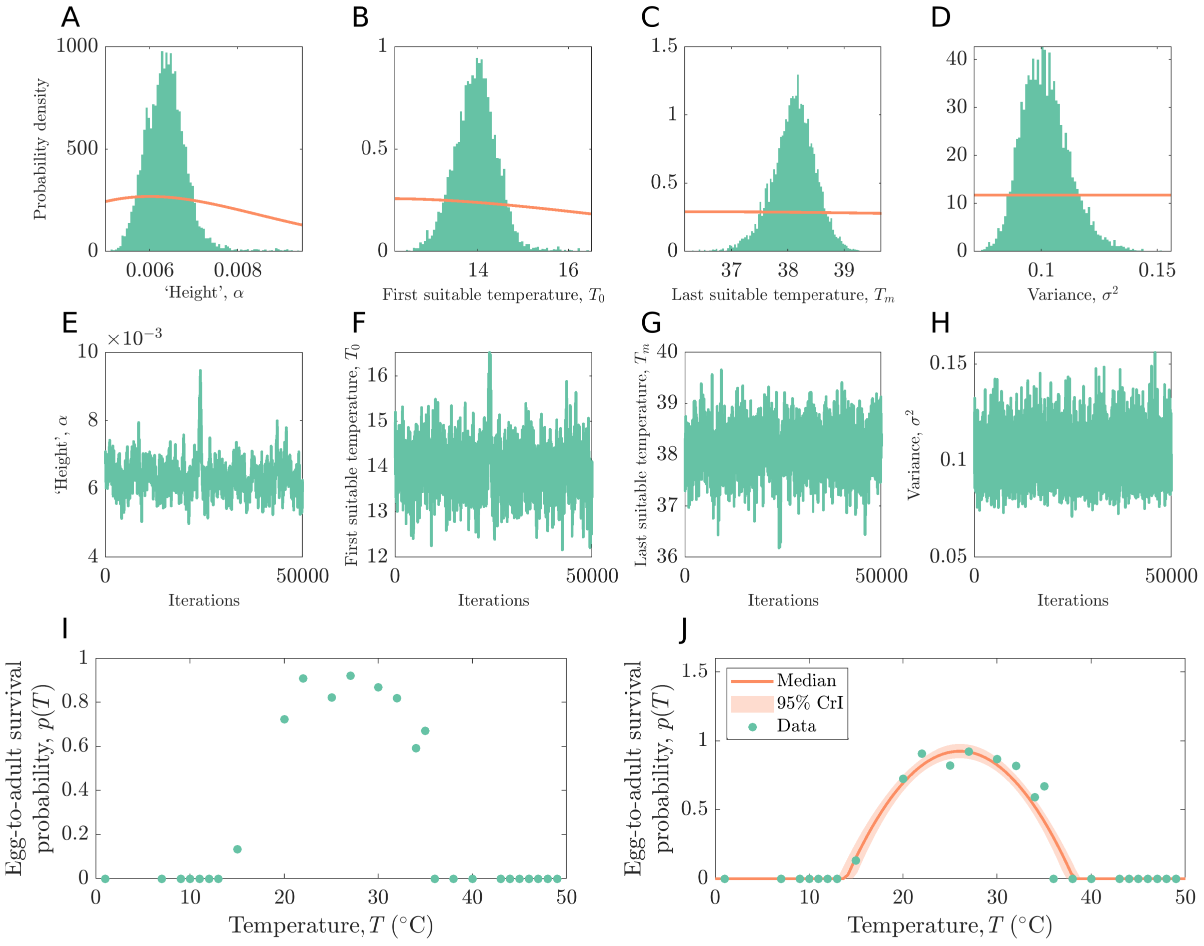


**Fig S4. The dependence of the *Ae. aegypti* egg-to-adult survival probability ($p(T)$) on temperature.** A-D. Prior (red) and posterior (green) distributions for each of the fitted sub-parameters ($\alpha$, $T_0$, $T_m$ and $\sigma^2$). To allow the posterior distribution to be seen clearly, x-axes limits are restricted to the minimum and maximum values in the posterior. E-H. Trace plots corresponding to the posterior distributions shown in panels A-D. 100,000 steps were run in the MCMC chain, with the first 50,000 discarded as burn-in (acceptance rate: 0.2267). Five chains were run to compute the Gelman-Rubin statistic (which was 1.0048, 1.0031, 1.0024 and 1.0004 for $\alpha$, $T_0$, $T_m$ and $\sigma^2$, respectively); the trace plots in panels E-H are from the first chain. I. Data describing the *Ae. aegypti* egg-to-adult survival probability as a function of temperature. J. Quadratic equation fit to the data in panel I (data – green; median fit – red; 95% equal-tailed credible interval – shaded region). Fitted values were constrained to lie between zero and one so that $p(T)$ represents a valid probability (MCMC steps corresponding to values outside of this range were discarded).

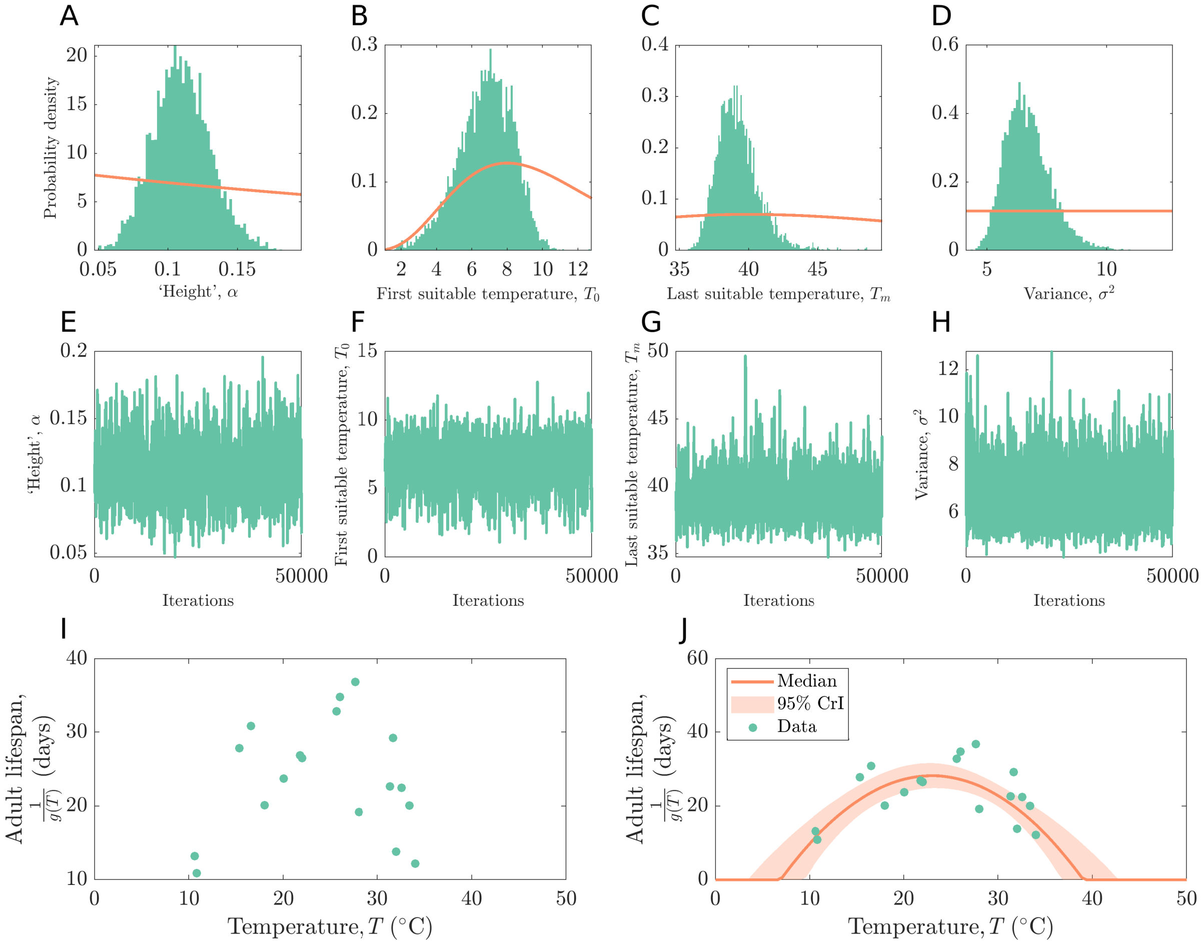


**Fig S5. The dependence of the *Ae. aegypti* adult lifespan ($1/g(T)$) on temperature.** A-D. Prior (red) and posterior (green) distributions for each of the fitted sub-parameters ($\alpha$, $T_0$, $T_m$ and $\sigma^2$). To allow the posterior distribution to be seen clearly, x-axes limits are restricted to the minimum and maximum values in the posterior. E-H. Trace plots corresponding to the posterior distributions shown in panels A-D. 100,000 steps were run in the MCMC chain, with the first 50,000 discarded as burn-in (acceptance rate: 0.2141). Five chains were run to compute the Gelman-Rubin statistic (which was 1.0008, 1.0003, 1.0014 and 1.0001 for $\alpha$, $T_0$, $T_m$ and $\sigma^2$, respectively); the trace plots in panels E-H are from the first chain. I. Data describing the *Ae. aegypti* adult lifespan as a function of temperature. J. Quadratic equation fit to the data in panel I (data – green; median fit – red; 95% equal-tailed credible interval – shaded region).

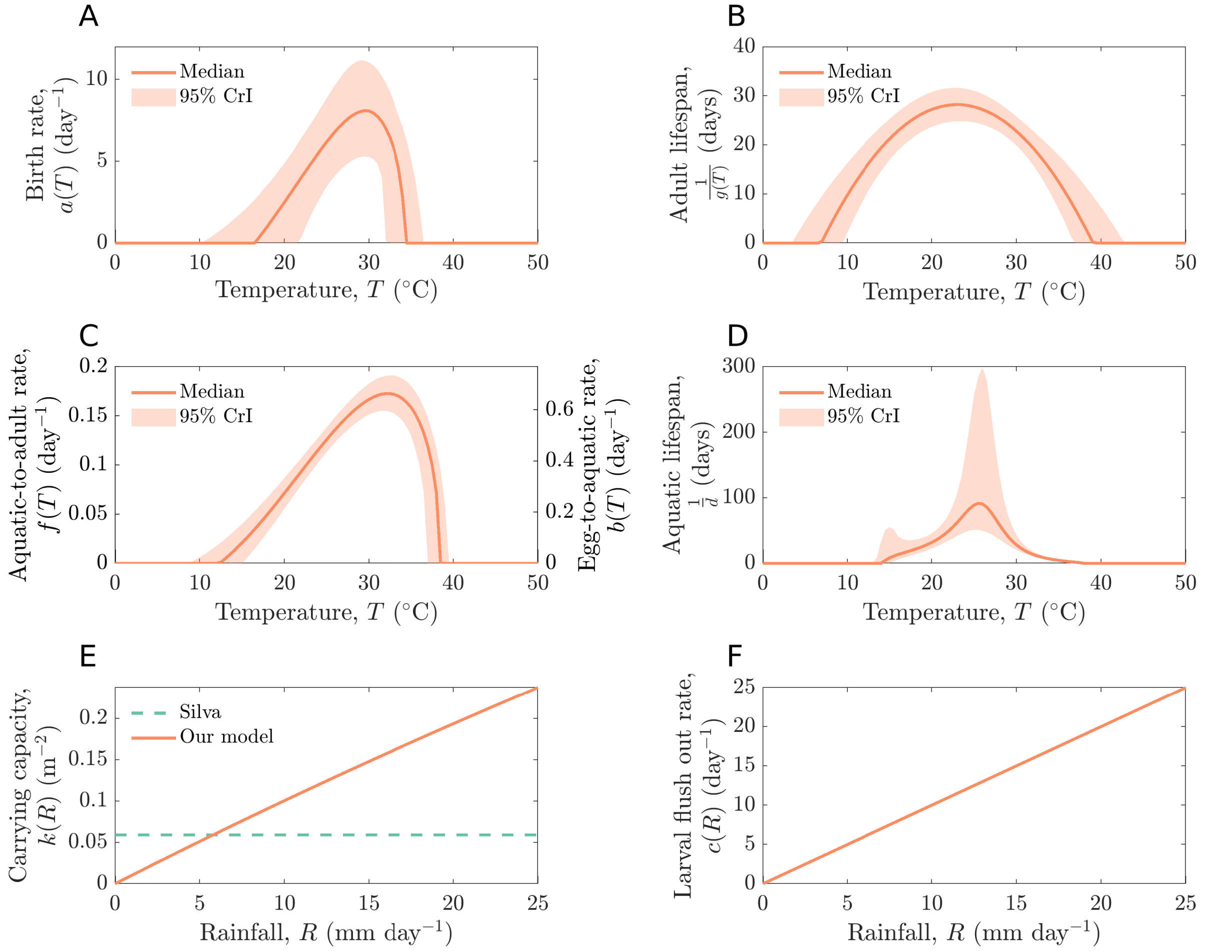


**Fig S6. The dependence of the parameters of the ecological model on temperature and rainfall.** Posterior distributions are shown for temperature-dependent parameters (median fit – red; 95% equal-tailed credible interval – shaded region): A. Birth rate ($a(T)$); B. Adult lifespan ($1/g(T)$); C. Aquatic-to-adult development rate (left y-axis, $f(T)$) and egg-to-aquatic development rate (right y-axis, $b(T)$); D. Aquatic stage lifespan ($1/d(T)$). Rainfall-dependent parameter responses (red) for: E. Aquatic stage carrying capacity per unit area ($k(R)$); F. Larval flush out rate ($c(R)$). In panel E, the aquatic stage carrying capacity estimated in a previous study[2] is also plotted for comparison (green dotted); in that study location (Nova Iguaçu, Brazil), the average rainfall is 4.89mm per day, which is comparable with the output from our model.

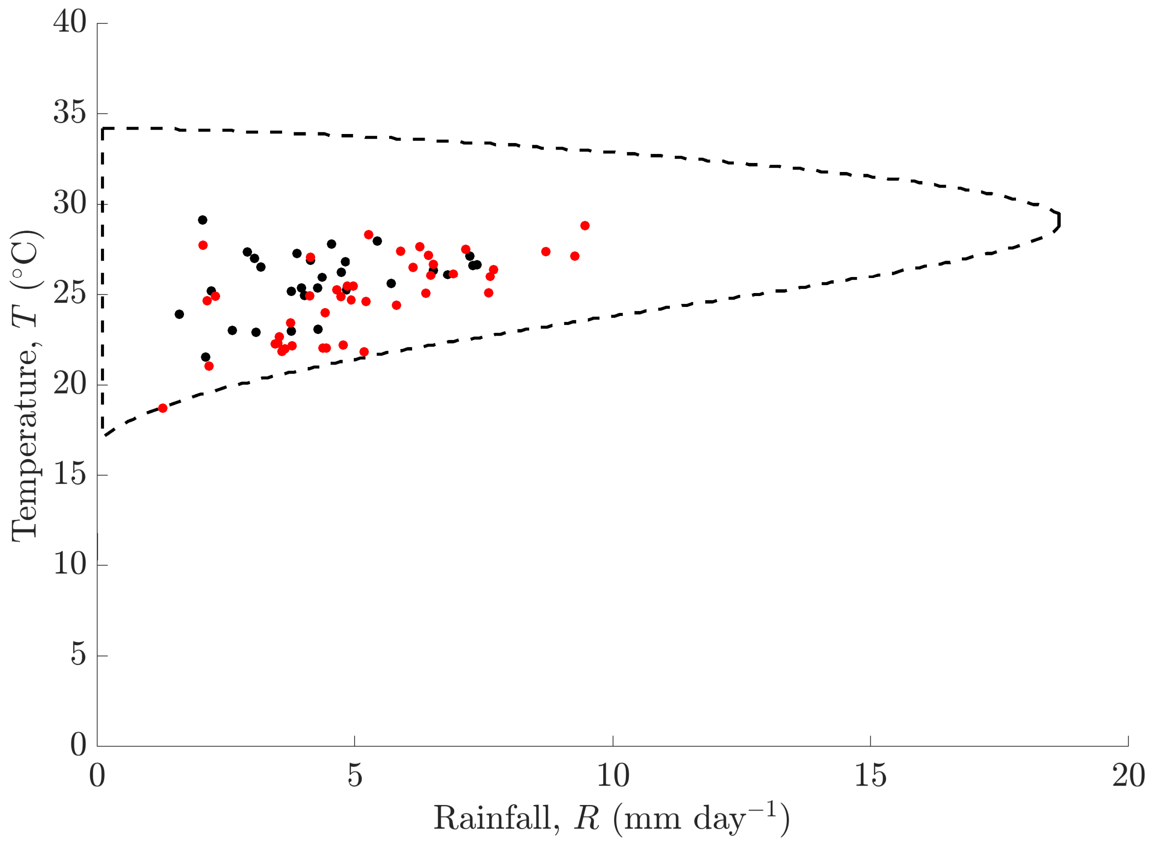


**Figure S7. Comparison of the ecological niche derived from our model and real-world data.** The black dotted line is the 50$^{th}$ percentile ecological niche (Fig 1C in the main text). Black dots indicate the mean temperature and rainfall in 29 countries with confirmed *Ae. aegypti* populations (locations were reported in Kraemer *et al.*[8], and temperature and rainfall values were averaged values across 2015 extracted from the World Bank's Climate Change Knowledge Portal[9]). Red dots indicate temperature and rainfall values for 48 locations that have experienced outbreaks of dengue virus disease (locations were reported by Liu *et al.*[10], and temperature and rainfall values for each outbreak are mean values across the period of the outbreak extracted from the World Bank's Climate Change Knowledge Portal[9] and the National Centers for Environmental Information's Climate Data Online tool[11]).

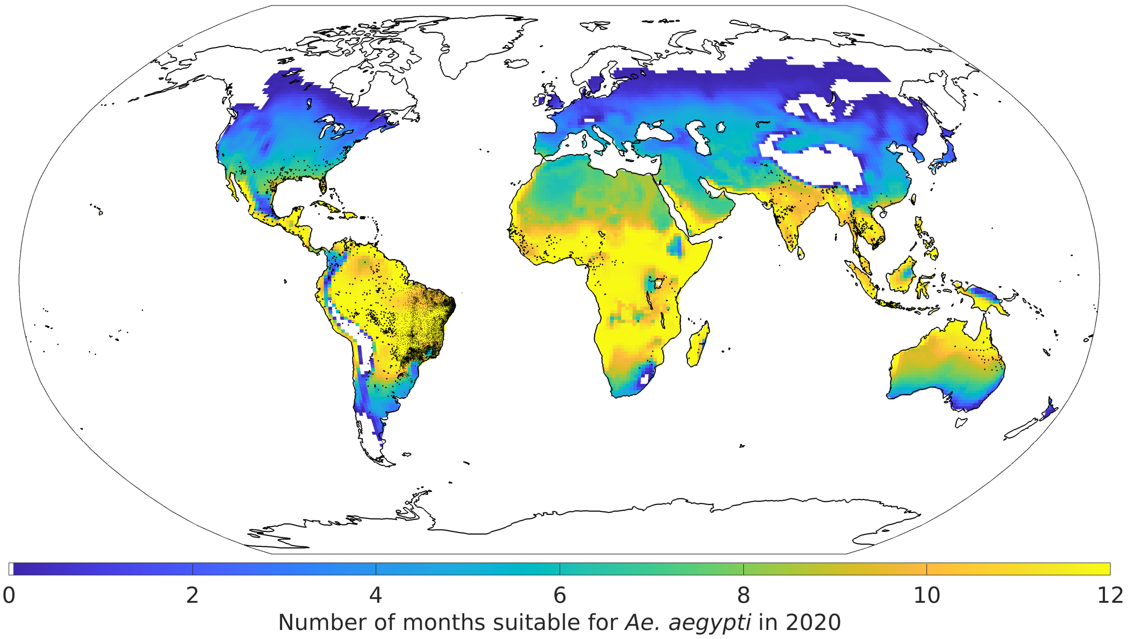


**Figure S8. Comparison of model-predicted global suitability for *Ae. aegypti* and known locations with *Ae. aegypti* in 2020.** The number of months that are predicted to be suitable for *Ae. aegypti* in different locations globally in 2020. These results were obtained first for each CESM simulation individually, and then averaged across all CESM simulations. Black dots indicate locations with recorded *Ae. aegypti* populations.[8]

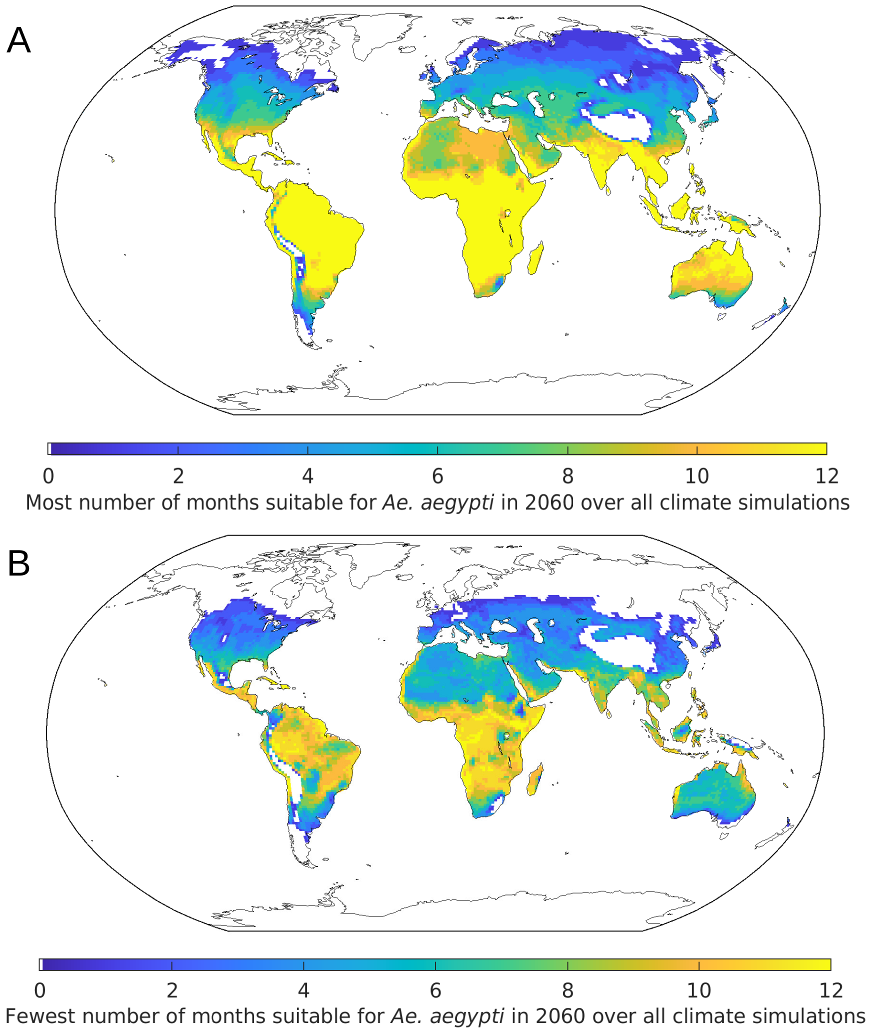


**Figure S9. The impact of climate variability on projected suitability for *Ae. aegypti* in 2060 in different locations.** A. The maximum number of months that are projected to be suitable for *Ae. aegypti* in the year 2060. B. The minimum number of months that are projected to be suitable for *Ae. aegypti* in the year 2060. In both panels, for each latitude-longitude value, the CESM projection corresponding to the most (panel A) or fewest (panel B) number of months that are suitable for *Ae. aegypti* in the year 2060 is chosen. This figure is analogous to Fig 3 in the main text, but for the year 2060 rather than 2100.

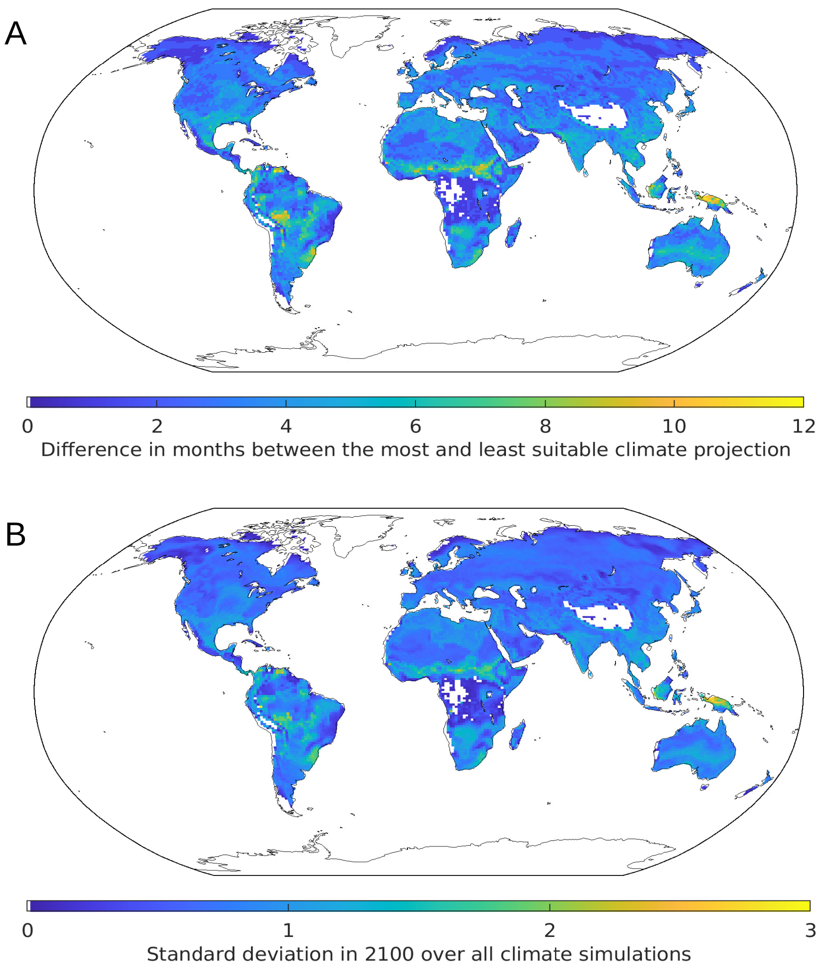


**Figure S10. Geographical variability in the impact of natural climate variability on climate suitability for *Ae. aegypti* in different locations.** A. Difference between the minimum and maximum number of months of 2100 in each location that are projected to be suitable for *Ae. aegypti*. This plot shows the difference between Figs 3A and 3B in the main text. B. The standard deviation in the number of months that are projected to be suitable for *Ae. aegypti* in the year 2100 across the CESM projections. Values shown in panel B are computed from the full range of 100 CESM projections.

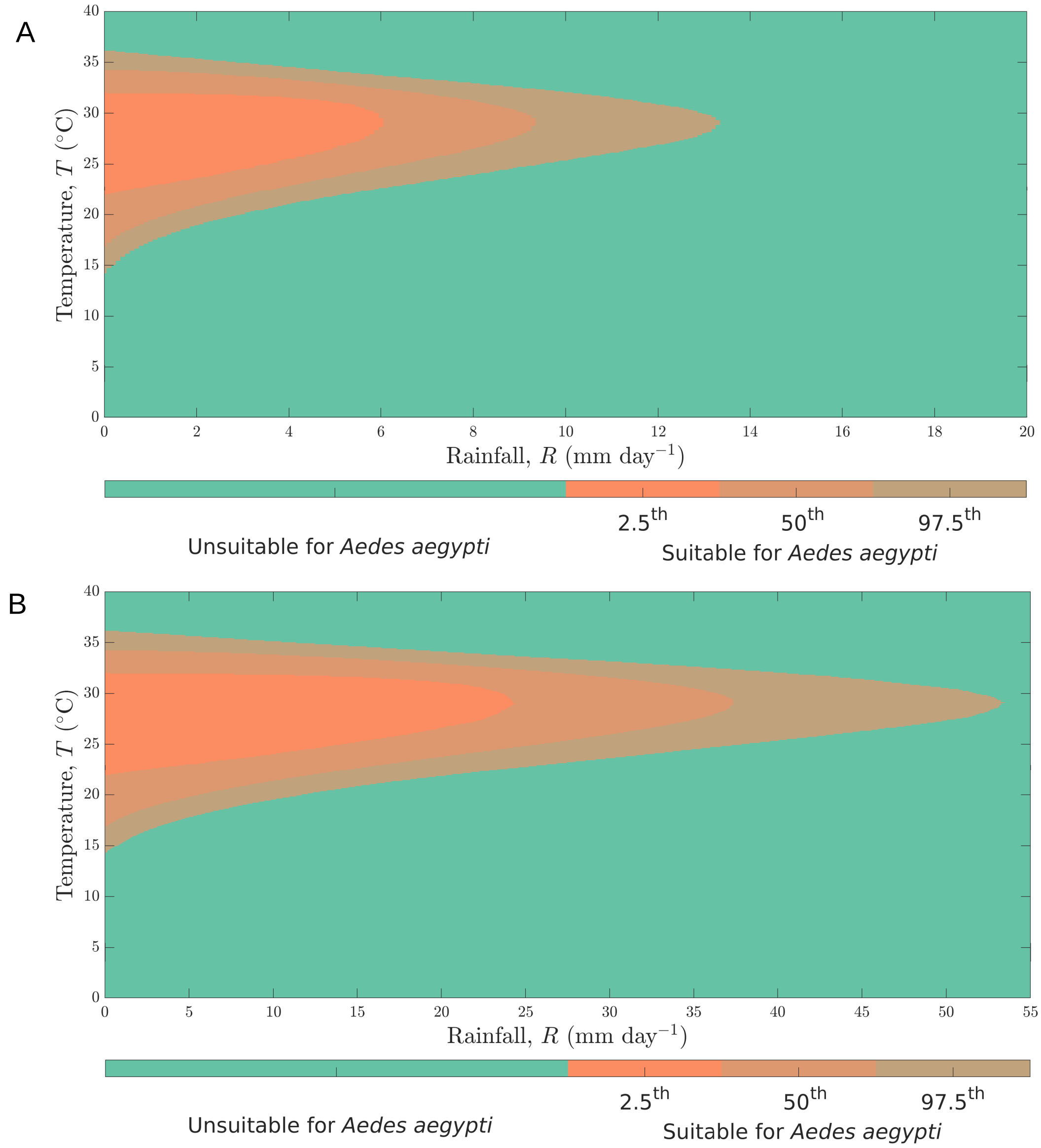


**Fig S11. Sensitivity of the ecological niche to the relationship between rainfall and the probability that aquatic stage individuals are washed away.** A. The ecological niche derived from the ecological model when aquatic stage individuals are washed away at a faster rate than assumed in our main analyses ($c(R) = 2R$). B. The ecological niche derived from the ecological model when aquatic stage individuals are washed away at a slower rate than assumed in our main analyses ($c(R) = \frac{1}{2}R$). Uncertainty in the ecological niche is represented by different shades of orange and arises due to uncertainty in the parameter estimates of the ecological model.

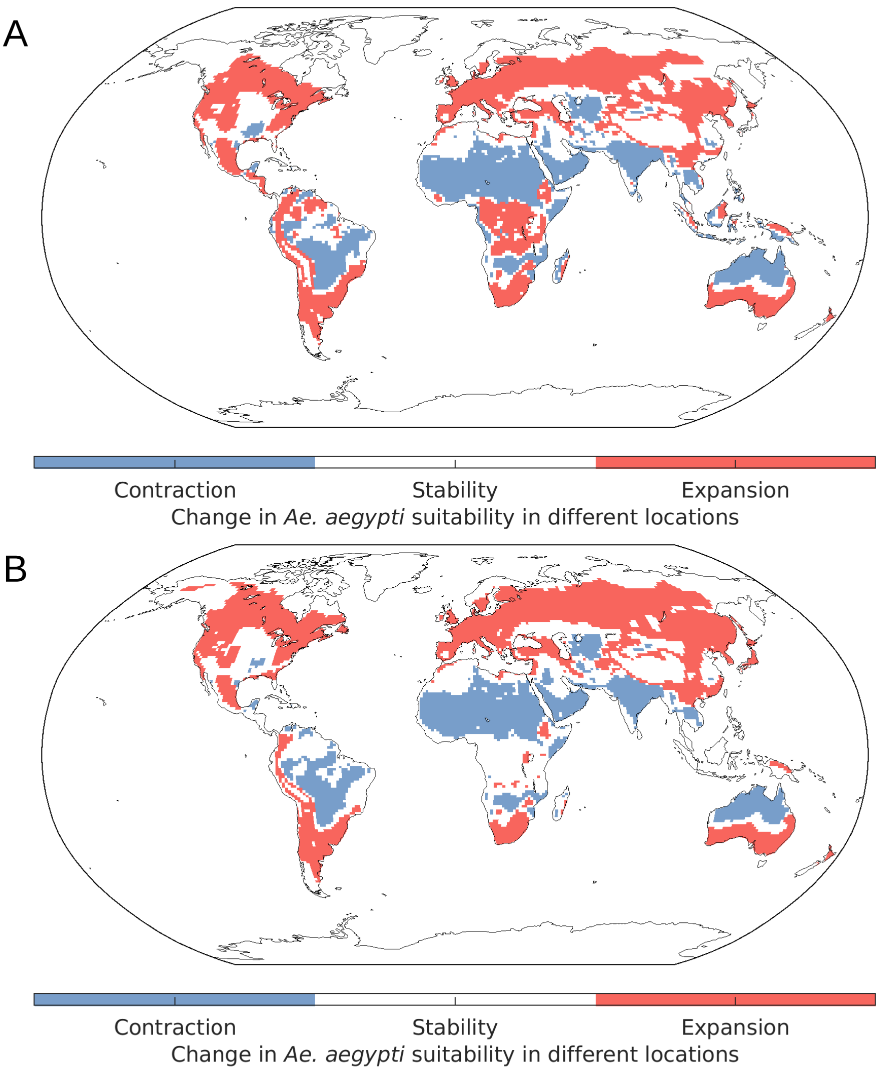


**Fig S12. Locations that are expected to see an increase or decrease in the suitability of climatic conditions for *Ae. aegypti*, for different ecological niches.** Results shown here are analogous to those in Fig 2 in the main text, but for: A. The ecological niche shown in Fig S11A ($c(R) = 2R$). B. The ecological niche shown in Fig S11B ($c(R) = \frac{1}{2}R$). Locations in which the number of months that are suitable for *Ae. aegypti* increases by more than one in 2100 compared to 2020 are shown in red. Locations with a corresponding decrease are shown in blue. In each panel, the results were obtained by first calculating the change in the number of suitable months for each CESM projection individually, and then averaging across all projections.

#### References

1 Mordecai EA, Cohen JM, Evans MV, *et al.* Detecting the impact of temperature on transmission of Zika, dengue, and chikungunya using mechanistic models. *PLoS Negl Trop Dis* 2017; **27**: e0005568.

2 Silva MR, Lugão PHG, Chapiro G. Modeling and simulation of the spatial population dynamics of the *Aedes aegypti* mosquito with an insecticide application. *Parasites Vectors* 2020; **13**: 550.

3 Roberts GO, Rosenthal JS. Optimal scaling for various Metropolis-Hastings algorithms. *Statist Sci* 2001; **16**: 351–67.

4 Tompkins AM, Ermert V. A regional-scale, high resolution dynamical malaria model that accounts for population density, climate and surface hydrology. *Malar J* 2013; **12**: 65.

5 Kittayapong P, Kaeothaisong N, Ninphanomchai S, Limohpasmanee W. Combined sterile insect technique and incompatible insect technique: sex separation and quality of sterile *Aedes aegypti* male mosquitoes released in a pilot population suppression trial in Thailand. *Parasites Vectors* 2018; **11**: 657.

6 Parham PE, Michael E. Modeling the effects of weather and climate change on malaria transmission. *Environ Health Perspect* 2010; **118**: 620–6.

7 National Center for Atmospheric Research. The Climate Data Guide: Global surface temperatures (BEST: Berkeley Earth Surface Temperatures). 2023 www.climatedataguide.ucar.edu/climate-data/global-surface-temperatures-best-berkeley-earth-surface-temperatures.

8 Kraemer MUG, Sinka ME, Duda KA, *et al.* The global distribution of the arbovirus vectors *Aedes aegypti* and *Ae. albopictus*. *eLife* 2015; **4**: e08347–e08347.

9 World Bank Group. Climate Change Knowledge Portal (CCKP). 2022 https://climateknowledgeportal.worldbank.org/.

10 Liu Y, Lillepold K, Semenza JC, Tozan Y, Quam MBM, Rocklöv J. Reviewing estimates of the basic reproduction number for dengue, Zika and chikungunya across global climate zones. *Environment Res* 2020; **182**: 109114.

11 National Oceanic and Atmospheric Administration. National Centers for Environmental Information: Climate Data Online. 2023. www.ncdc.noaa.gov/cdo-web/.
